# The Microbiome-Inflammation Axis in Pediatric Cardiac Surgery: Decoding Functional Bacterial Responses

**DOI:** 10.64898/2026.07.01.26357082

**Authors:** Haowen Qiu, Monalisha Elango, Jean Jack Riethoven, Gleb Haynatzki, Ali Ibrahimiye, Camille Hancock Friesen, Mabruka Alfaidi, Ram K Subramanyan, Jeffrey D Salomon

**Author notes:** **Funding:** 1. American Heart Association – 23CDA1045271, 7272 Grenville Ave, Dallas, TX 75231 2. National Institute for General Medical Science – 1P20GM152326-01, P20GM103427-19 & U54GM115458, 45 Center Drive, Bethesda, MD 20892 3. Child Health Research Institute, University of Nebraska Medical Center, 45^208^ & Emile St, Omaha, NE 68198. **Disclosures:** There are no disclosures or any industry relationships for any authors. **Address for Correspondence:** Jeffrey D Salomon, 8200 Dodge St. Omaha, NE, 68114.

## Abstract

**Background:** Gut injury after pediatric cardiac surgery remains an ongoing challenge, resulting in increased morbidity and mortality for children with congenital heart disease (CHD) and a significant burden on the healthcare system. It remains unclear what the driving forces are that result in this pro-inflammatory state following pediatric cardiac surgery with cardiopulmonary bypass. Understanding key components involved in the gut composition, gut barrier function, and systemic inflammation in children with CHD after cardiac surgery is vital to improve outcomes.

**Methods:** A prospective study of patients aged 0-5 years with CHD undergoing cardiac surgery (CPB group) or non-CHD undergoing non-cardiac surgery (Comparison group). We collected pre-operative and post-operative stool and plasma to evaluate the microbiome, metabolites, markers of gut barrier function, and inflammatory cytokines. Clinical variables were collected to evaluate markers of inflammation. These variables were compared between the two groups to evaluate signatures and develop unique biomarker profiles.

**Results:** We enrolled 62 patients (CPB, n=46; Comp, n=16). CPB patients had increased pro-inflammatory microbiota and reduced diversity metrics pre-operatively, which were exacerbated post-operatively. The CPB group also had increased pro-inflammatory eicosanoids and reduced gut and heart protective short-chain fatty acids versus the Comparison group. The CPB group had increased pro-inflammatory and reduced anti-inflammatory cytokines post-operatively. The CPB group also had increased markers of gut barrier dysfunction versus the Comparison group. Mediation analysis showed the microbial functional shift was associated with increased PGE2 and reduced butyric acid in the CPB group, associated with increased cytokines and clinical markers of inflammation post-operatively.

**Conclusion:** We demonstrate unique gut microbial and metabolites profiles associated with gut permeability and systemic inflammation in children with CHD undergoing cardiac surgery highlighting a unique microbiome-inflammation axis in this patient population. Further studies to evaluate causal links with these profiles will identify potential targets to improve outcomes for these patients.

## Introduction

Congenital heart disease (CHD) stands as the most prevalent birth defect, impacting over 40,000 newborns each year^1^. The vast majority of affected infants require surgical repair, and nearly one in four presents with critical CHD, demanding immediate surgical intervention within the first weeks of life. These delicate procedures rely on cardiopulmonary bypass (CPB) — a life-saving technique that, however, initiates a profound systemic inflammatory response syndrome (SIRS)^2^. This inflammation cascade is influenced by the gut microbiome and its metabolites, which can amplify immune activation both locally and throughout the body.

The gut microbiome serves as a cornerstone of systemic health and dysbiosis, characterized by increased pro-inflammatory and decreased beneficial bacteria, which heightens inflammation and alters gene expression. Gut complications in children with CHD occur in roughly 33% of patients^3-6^. Children with CHD exhibit notable microbial imbalances, including higher levels of pro-inflammatory bacteria, elevated inflammatory metabolites, gut barrier dysfunction after CPB, and exaggerated systemic immune activation^6-10^. These metabolic disturbances, marked by an increase in harmful compounds and a depletion of protective ones, have been linked to worse clinical outcomes^11,12^, though the mechanisms remain unclear.

Among these mediators, inflammatory lipid molecules known as eicosanoids, encompassing prostanoids, leukotrienes, and lipoxins, play a pivotal role. Eicosanoids drive cytokine release, mobilize immune cells, and fuel inflammatory cascades within both the intestine and circulation^13-16^. Altered gut microbial communities in children with CHD increase intestinal eicosanoid production, a process further exacerbated by CPB^7,17,18^. The result: widespread inflammation, beginning in the gut and echoing through the entire body.

In contrast, short-chain fatty acids (SCFAs), which are beneficial metabolites generated by bacteria such as *Prevotella, Clostridia, Roseburia, Lachnospiraceae,* and *Ruminococcus,* provide gut and heart protection^19-22^. SCFA nourish the intestinal lining, stimulate regulatory T-cell activity, regulate calcium and blood pressure, and support cardiac healing^23,24^. Yet, children with CHD have SCFA-producing bacteria, an imbalance that correlates with serious gut complications, including feeding intolerance and necrotizing enterocolitis^6^.

While the gut-heart connection has been well-documented in adults with cardiovascular disease, pediatric research remains scarce. Considering the frequency and severity of gut-related complications among children with CHD, deciphering the interplay between microbiome, metabolites, and inflammation is essential. By mapping gut microbial and metabolite profiles, markers of gut barrier dysfunction, cytokines, and inflammatory gene expression in CHD patients undergoing cardiac surgery and contrasting them with those in non-CHD children undergoing non-cardiac surgeries, our study seeks to pinpoint markers. This may uncover novel therapeutic targets within the gut–heart axis to improve post-operative recovery and long-term outcomes.

## Methods

### Patient Enrollment and Surgery

This prospective observational cohort study, approved by the institutional review board of the University of Nebraska Medical Center, was conducted from January 2023 through June 2024. Informed consent was obtained from the legal guardians of all subjects, and patients were enrolled through Children’s Nebraska in Omaha, NE (IRB #0070-20-FB). This study conformed to the principles outlined in the Declaration of Helsinki. Patients with CHD undergoing cardiac surgery with CPB and patients without CHD undergoing non-cardiac surgery were enrolled in the experimental and comparison arms, respectively. The comparison arm accounted for shared perioperative variables, including *nothing per os* (NPO) time, anesthetics, perioperative antibiotics, mechanical ventilation, and surgical duration. Inclusion criteria were children aged 0–10 years undergoing either cardiac surgery with CPB or non-cardiac surgery with expected surgical duration greater than 90 minutes and hospital length of stay ≥2 two days. Exclusion criteria included prior surgery in the past three months, antibiotic use within eight weeks, gastrointestinal pathology or intestinal surgery (excluding gastrostomy tube), continuous enteral feeding, renal disease requiring dialysis, or liver disease with transaminases two-times the upper limit of normal.

Surgical procedures and anesthetic regimens followed the standard of care. Patients received perioperative and post-operative cefazolin unless a documented penicillin allergy or methicillin-resistant Staphylococcus aureus was present. Blood prime was used for CPB patients weighing less than 10 kg, and saline prime for all others. Modified ultrafiltration and cell-saver techniques minimized exogenous blood exposure. All surgeries requiring CPB and cross-clamp employed dual aortic cannulation, preserving splanchnic perfusion throughout CPB. Neither selective cerebral perfusion nor deep hypothermic circulatory arrest was performed in this cohort.

### Stool and Blood Preparation

Stool was collected prior to surgery either from a spontaneous bowel movement using the Easy Sampler® collection kit (ALPCO Diagnostics, Cat#58-EZSampler) and stored at −17 to −20°C until the morning of surgery, or via a sterile FecalSwab™ (Copan Diagnostics, Cat#4CO24S) collected in the operating room. The post-operative stool sample was obtained from the first post-operative bowel movement. All stool samples were transferred and stored at −80°C. DNA was extracted using the Qiagen PowerFecal DNA Extraction Kit and quantified using the NanoDrop™ One (Thermo Fischer Scientific). Blood samples were collected pre-operatively, one hour post-operatively, and on post-operative days (POD) 1–3. Plasma was isolated and stored at −80°C per our previously published protocol^6,7,18^. Markers of EBD and cytokines were measured at all 5 time points, while plasma SCFA and Eicosanoids were measured pre-operatively and at the 1-hr post-op time point.

### Library Preparation, Sequencing, Bioinformatics, and Microbiome Analysis

Shotgun metagenomic sequencing libraries were prepared from extracted fecal DNA using the Illumina DNA Prep kit and sequenced on the Illumina NovaSeq 6000 platform, generating 2 × 150 bp paired-end reads. Raw sequencing reads were processed through the bioBakery 3 pipeline^25^. Taxonomic profiling was performed using MetaPhlAn 4, generating relative abundance tables and was imported into R (v4.3) for diversity analysis^26^. Functional profiling was performed using HUMAnN 3, which reconstructs microbial metabolic pathways by mapping quality-filtered reads to the MetaCyc pathway database via species-stratified, pangenome-level gene mapping. Alpha diversity was calculated using multiple metrics, and differences between groups were assessed using Wilcoxon rank-sum tests for unpaired comparisons and Wilcoxon signed-rank tests for paired comparisons. Beta diversity was assessed using Bray-Curtis dissimilarity and Jaccard distance matrices. Differences in composition were tested using permutational multivariate analysis of variance (PERMANOVA). Principal coordinates analysis (PCoA) was used to visualize differences. PERMANOVA models were fit for the overall cohort and within the CPB group for age, cyanotic status, and clinical outcome. Differential abundance was performed using the corncob package (v0.4) in R^27^. Differential pathway abundance was performed using DESeq2. Benjamini-Hochberg (BH) false discovery rate (FDR) correction was applied, and taxa were considered differentially abundant at FDR < 0.05.

### ELISA, Immunoassay Multiplex, and Metabolomics

Claudin-2 and claudin-3 (MyBiosource, Cat# MBS165353, Cat# MBS160582) and intestinal fatty acid binding protein (FABP2; Sigma-Aldrich, Cat# RAB0537-1KT) were measured in plasma from CPB patients by ELISA following previously published protocols. Plasma cytokines were quantified using the ProcartaPlex magnetic bead-based immunoassay kit (ThermoFisher Scientific, Cat# PPX-09-MXGZGAK) read on a Luminex 100/200 system^6^. A panel of short-chain fatty acids (SCFAs) and a panel of eicosanoids were profiled from stool and plasma with quantification using ultra-performance liquid chromatography-tandem mass spectrometry (UPLC-MS/MS) on a Shimadzu 8060NX mass spectrometer (Shimadzu Scientific Instruments, Columbia, MD), per previously published protocols^6,7,18^. SCFA and eicosanoids levels were determined using LabSolution software (version 5.99). Analytes that did not reach quantifiable levels were excluded from the analysis.

### Biomarker and Clinical Statistical Analysis

Continuous variables were reported as either mean ± SD or median (interquartile range), depending on excessive skewness or outliers in the distributions. Categorical data were documented as counts and percentages. We primarily use the chi-square test to analyze the frequency of a categorical variable; Fisher’s Exact test was used when the frequency was smaller. The Shapiro-Wilk method assessed data normality and either an independent t-test or a Wilcoxon rank-sum test was conducted. For normally distributed biomarker variables, two-way ANOVA or mixed-effects analysis was performed. Post hoc test using the Tukey test for multiple comparisons. A two-sided p-value of 0.05 was considered significant. Biomarker analysis was performed using GraphPad Prism (version 9.5.0). All statistical analyses have been performed using the SAS/ACCESS® 9.4 Interface to ADABAS software.

### Canonical Correlation and Mediation Analysis

Regularized canonical correlation analysis (rCCA) was performed using the shrinkage method implemented in the mixOmics package (v6.24) in R^28^. Analyses were performed for stool SCFAs, stool eicosanoids, plasma SCFAs, plasma eicosanoids, plasma cytokines, and plasma EBD markers. Clustered image maps (CIM) were generated from the rCCA correlation matrices. Pairwise Spearman rank correlations were computed between taxon-metabolite pairs within each panel, with BH correction applied across all tests.

We performed mediation analysis using the MODIMA (Multivariate Omnibus Distance Mediation Analysis) framework^29^ implemented via the energy package (v1.7) and the ade4 (v1.7) package^30^. MODIMA was structured with a distance-based framework, evaluating the mediation path Exposure (X) → Microbiome (M) → Response (Y), where the microbiome is represented as a Jensen-Shannon divergence (JSD) distance matrix computed from species-level relative abundance profiles. The Benjamini-Hochberg (BH) correction was applied to metabolite models. Results were considered significant at FDR < 0.05.

## Results

### Patient Characteristics

A total of 46 CPB patients and 16 Comparison patients were included in the analysis. Demographic characteristics for both groups and CPB subgroups are listed in **Table 1**. There were no significant differences between the CPB and Comparison groups in age, weight, surgical time, antibiotic use, or NPO time. A significant difference in sex distribution was observed, with 12 females (26.1%) in the CPB group and 11 females (73.3%) in the Comparison group (p < 0.05). Diagnoses and surgeries performed for the CPB group are listed in **Supplemental Table 1**. Comparison group surgeries consisted of cranial vault reconstruction (n=10), posterior fossa tumor resection (n=4), and open hip reduction (n=2).

Significant differences in post-operative clinical and laboratory values between the CPB and Comparison groups are shown in Table 2. CPB patients had markedly longer hospital length of stay (CICU: 11.6 vs. 2.1 days; Hospital: 16.9 vs. 5.1 days), higher vasoactive inotrope score (VIS: 7.0 vs. 0.0), and greater requirement for post-operative nasogastric feeds (60.9% vs. 6.7%). CPB patients also had elevated post-operative BUN (21.8 vs. 9.5 mg/dL), creatinine (0.6 vs. 0.22 mg/dL), and PELOD score (6.3 vs. 2.7), with lower post-operative hematocrit (36.8 vs. 29.7%). Clinical variables are reported in **Table 2**.

### Microbiome Analysis

#### Abundance and Diversity

Whole genome shotgun sequencing was performed on pre-operative and post-op stool samples. Relative abundance bar plots at the phylum level showed that Bacillota (Firmicutes) and Bacteroidota (Bacteroidetes) dominated both groups preoperatively. The CPB group showed fewer distinct phyla, reduced relative abundance of Bacillota, and increased relative abundance of Pseudomonadota (Proteobacteria). Both groups showed an increased relative abundance of Verrucomicrobiota and Bacteroidota post-surgery, although the CPB group also had an increase in Pseudomonadota and further reduction in Bacillota (**Figure 1A**).

**Figure 1.**
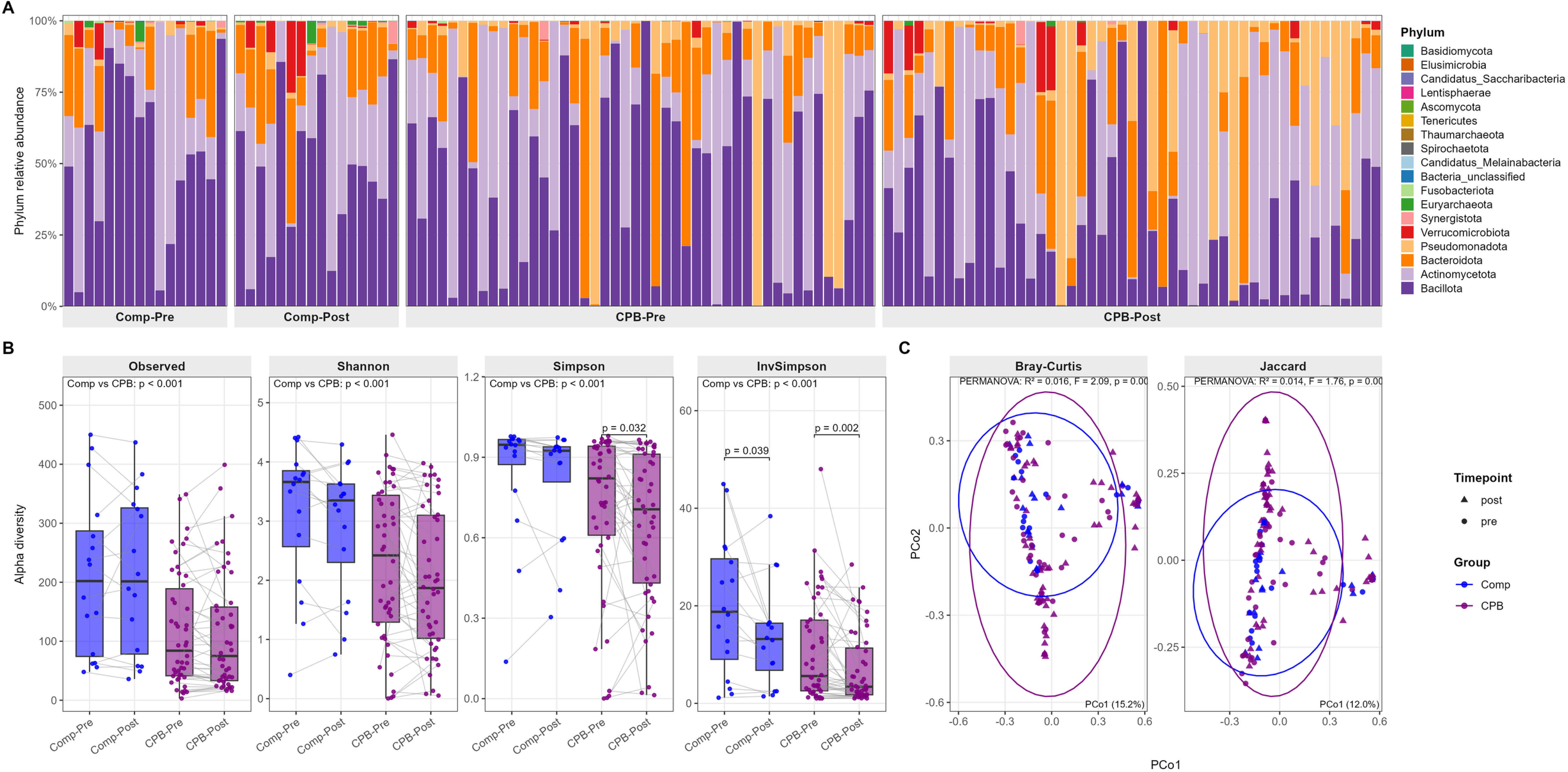
Gut microbiome composition and diversity in the CPB group versus the Comparison Group. Stool samples were collected pre-operatively and post-operatively from CPB patients and a non-cardiac surgical comparison group (Comp) and profiled by whole-genome shotgun metagenomic sequencing. (A) Phylum-level relative abundance per sample, faceted by group and timepoint (Control-Pre, Control-Post, CPB-Pre, CPB-Post). (B) Alpha diversity for four metrics: Observed richness, Shannon, Simpson, and Inverse Simpson indices. Grey lines connect paired pre- and post-operative samples from the same patient. (C) Principal coordinates analysis (PCoA) of beta diversity using Bray-Curtis (left) and Jaccard (right) dissimilarity matrices computed on relative-abundance-transformed counts. Color denotes group (blue = Control, purple = CPB) and shape denotes timepoint (circle = pre, triangle = post). 95% confidence ellipses are drawn per group. PERMANOVA results for the overall group effect are annotated within each panel.

Alpha diversity was reduced in the CPB group versus the Comp group (Observed: p<0.001; Shannon: p<0.001; Simpson: p=0.001; Fisher: p<0.001) and when stratified by timepoint in both pre-op and post-op (**Figure 1B**). Paired Wilcoxon signed-rank tests within the CPB group revealed a post-op decline in community evenness, with Shannon (p=0.033), Simpson (p=0.032), and Inverse Simpson (p<0.001) indices decreasing between pre- and post-op time points. Richness metrics (Observed, Chao1, Fisher) did not change in either group. No pre- to post-op changes in alpha diversity were observed in the Comp group. PERMANOVA analysis demonstrated compositional differences between groups regardless of time point using Bray-Curtis (p=0.009) and Jaccard (p=0.011) distance matrices (**Figure 1C**). Beta diversity also differed between pre- and post-op time points (Bray-Curtis: p=0.001; Jaccard: p=0.001).

#### Differential Abundance and Functional Pathway Analysis

To identify specific taxa driving the observed compositional differences, differential abundance analysis was performed. At the post-operative time point, 25 genera and 34 families were depleted in the CPB group versus the Comparison group (FDR< 0.05), with no taxa enriched in the CPB group (**Figure 2A**). Depleted genera included *Blautia, Romboutsia, Prevotella, Christensenella,* and *Butyricimonas*. Depleted families included Lachnospiraceae, Oscillospiraceae, Eggerthellaceae, Christensenellaceae, Prevotellaceae, and Methanobacteriaceae. At the phylum level, Euryarchaeota were depleted in CPB patients. Pre-operatively, the CPB group showed depletion of one genus (*Blautia*) and five families, confirming dysbiosis prior to surgery in CHD.

**Figure 2.**
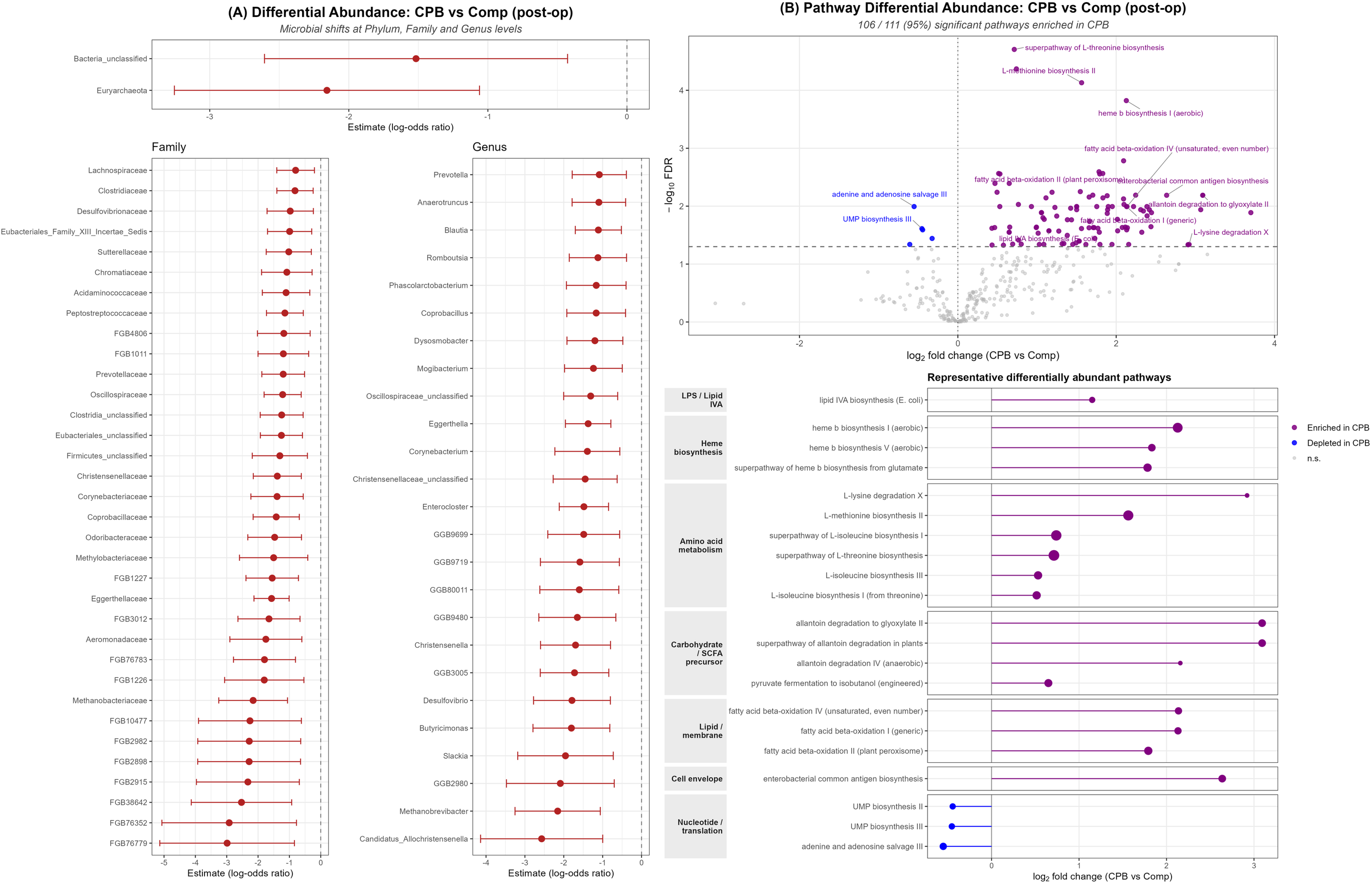
Differential abundance of gut taxa and microbial metabolic pathways in the CPB group versus the comparison group. Compared to a non-cardiac surgical comparison group, post-operatively. (A) Differential abundance analysis of gut microbial taxa between CPB and Comparison (Comp) patients at the post-operative timepoint. Each point shows the regression coefficient (log-odds ratio for relative abundance in CPB vs. Comp) with 95% confidence intervals, and only taxa that reach FDR significance are displayed. Top: Phylum level. Bottom left: Family level. Bottom right: Genus level. (B) Differential pathway abundance between CPB and Comparison patients at the post-operative time point. Top: Volcano plot of all tested pathways. The x-axis shows log₂ fold change (CPB vs Comp); purple points are pathways significantly enriched in CPB, blue points are significantly depleted in CPB, and grey points are non-significant. The horizontal dashed line marks the FDR = 0.05 cutoff. Selected pathways are labeled. Bottom: Curated lollipop plot of 21 representative significantly differentially abundant pathways, grouped by functional category. Point size is proportional to −log₁₀(FDR); color encodes direction (purple = enriched in CPB, blue = depleted in CPB).

Differential pathway abundance testing was performed using DESeq2 on HUMAnN 3-derived microbial metabolic pathways. Within the CPB group, 30 pathways were altered from pre- to post-surgery (FDR < 0.05), of which 27 were enriched post-operatively. Enriched pathways included L-threonine metabolism (log2FC = +2.57), L-lysine degradation X (log2FC = +2.74), three allantoin degradation routes, the pentose phosphate pathway (log2FC = +0.28), and the Entner-Doudoroff pathway III (log2FC = +9.90). Only three pathways were depleted post-operatively, all involved in cofactor biosynthesis: cobalamin (log2FC = −15.48), biotin (log2FC = −1.73), and molybdopterin (log2FC = −0.55). The near-complete loss of cobalamin biosynthesis paralleled the post-operative depletion of Methanobrevibacter and other Euryarchaeota. In contrast, only two pathways were altered in the comparison group between time points, demonstrating functional remodeling specific to cardiac surgery with CPB rather than to other perioperative variables. Post-operatively, 111 pathways differed between CPB and comparison patients, of which 106 were enriched in the CPB group, and only 5 were depleted (**Figure 2B**). Enriched pathways were dominated by functions characteristic of a Proteobacteria/Enterobacteriaceae-rich community, including amino acid biosynthesis (L-threonine, log2FC= +0.71; L-methionine, +1.56; L-isoleucine, +0.74), heme b biosynthesis I (+2.13), assimilatory sulfate reduction (+2.09), fatty acid β-oxidation (+1.79), and enterobacterial common antigen biosynthesis (+2.64). The five depleted pathways were limited to nucleotide metabolism and amino acid degradation (UMP biosynthesis, adenine/adenosine salvage, tRNA charging, and Stickland arginine degradation), all of which had modest effect sizes. Notably, lipid IVA biosynthesis — a vital step in lipopolysaccharide (endotoxin) synthesis — was enriched in the CPB group relative to comparison patients (log2FC= +1.15).

### Metabolomics

#### Short-Chain Fatty Acids

Of the eight SCFAs measured in stool, only valeric acid was lower in the CPB group compared to the Comparison group pre-operatively (p<0.001), while butyric acid trended toward a decrease (p=0.056) (**Figure 3A**). At the post-operative time point, propionic acid (p=0.008), butyric acid (p<0.001), and valeric acid (p<0.001) were all lower in the CPB group. Within the CPB group, seven of eight stool SCFAs were decreased post-operatively compared to pre-operatively (propionic acid: p<0.001; isobutyric acid: p=0.024; butyric acid: p<0.001; 2-methyl butyric acid: p<0.001; capric/isocapric acid: p=0.033), while isovaleric acid trended lower (p=0.100). No significant changes in any stool SCFA were observed in the Comparison group between timepoints.

**Figure 3.**
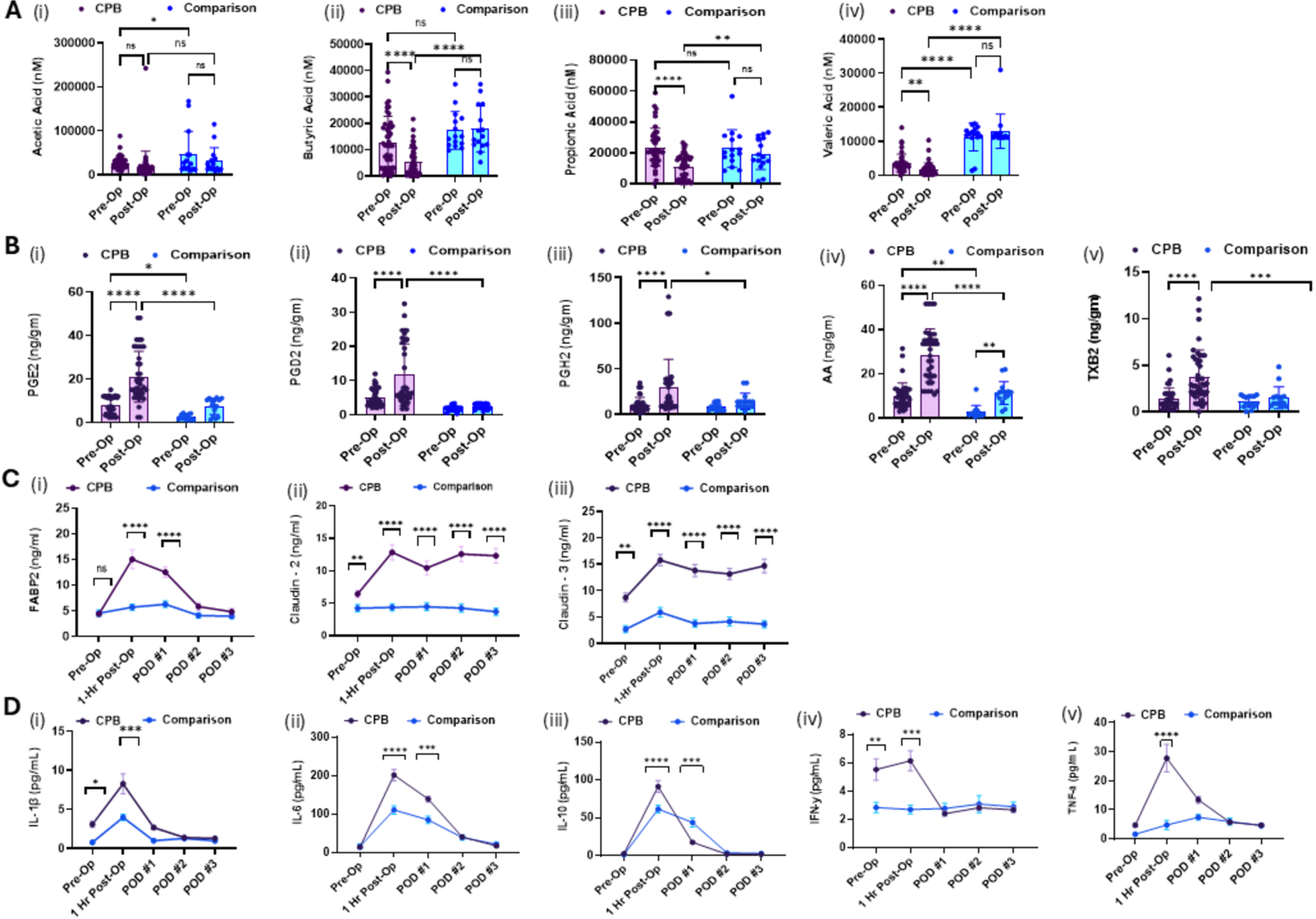
Changes in SCFA, eicosanoids, markers of EBD, and cytokines between the CPB group and the comparison group. Panel A shows reduced SCFA levels between pre-op and post-op time points in the CPB group compared with the Comp group. Panel B showed increased stool eicosanoids between pre-op and post-op time points in the CPB group versus the Comp group. Panel C shows increased circulating markers of EBD in the CPB group versus the Comp group. Panel D shows changes in plasma cytokines between pre-op and post-op time points in the CPB group versus the Comp group. Two-way ANOVA or Mixed effects analysis using Tukey test for multiple comparisons. Graphs displayed from GraphPad Prism. CPB, cardiopulmonary bypass; SCFA, short-chain fatty acids; PG, prostaglandin; AA, arachidonic acid; TXB, thromboxane; FABP, fatty acid binding protein; IL, interleukin; IFN, interferon; TNF, tumor necrosis factor; POD, post-op day. *p<0.05; **p<0.01; ***p<0.001; ****p<0.0001

Of the eight SCFAs measured in plasma, propionic acid (p=0.002), butyric acid (p<0.001), isovaleric acid (p=0.043), and valeric acid (p<0.001) were lower in the CPB group pre-operatively. At the 1-hr post-op time point, five SCFA were lower in the CPB group (propionic acid: p<0.001; butyric acid: p<0.001; isovaleric acid: p=0.032; valeric acid: p< 0.001; acetic acid: p=0.041). Within the CPB group, propionic acid (p<0.001), butyric acid (p<0.001), valeric acid (p<0.001), and acetic acid (p<0.001) were all decreased post-operatively; in the Comparison group, only valeric acid showed a significant pre-to-post decline (p=0.039).

#### Eicosanoids

Of the 22 eicosanoids measured in stool, over half were elevated in the CPB group compared to the Comparison group at the pre-operative timepoint, including PGE2 (p=0.003), PGD2 (p<0.001), PGE1 (p<0.001), arachidonic acid (AA; p<0.001), 11-HETE (p=0.003), 8-HETE (p=0.001), 15-Keto-PGE2 (p=0.013), 9(10)-DiHOME (p<0.001), 12(13)-DiHOME (p=0.002), 10-HDHA (p=0.011), 9(S)-HOTrE (p<0.001), 13(S)-HOTrE (p=0.016), 15-OxoETE (p=0.006), and 9-OxoODE (p<0.001), indicating that gut-level eicosanoid inflammation is present before surgery in CHD patients (**Figure 3B**). Post-operatively, the majority of stool eicosanoids were elevated in the CPB group, with the largest differences observed for PGE2 (p<0.001), AA (p<0.001), PGD2 (p<0.001), 9(S)-HOTrE (p<0.001), and 9-OxoODE (p<0.001). Within the CPB group, 11 stool eicosanoids increased from pre- to post-surgery including PGE2, AA, PGD2, 5-HETE, 13,14-DiOH-15-Keto-PGE2, PGH2, 10-HDHA, 11-HDHA, 12-HEPE, 9(S)-HOTrE, and 13(S)-HOTrE (all p≤0.05). In the Comparison group, PGE2, AA, PGE1, and PGD2 also increased post-surgery, though to a smaller extent than in the CPB group.

Of the 29 eicosanoids measured in plasma, few differed between groups at the pre-operative time point. PGD1 (p = 0.034) and 11-β-PGF2 (p=0.021) were increased in the CPB group, while LTB4 (p = 0.030) and docosahexaenoic acid (DHA; p=0.003) were lower. At the 1-hr post-op time point, 11 plasma eicosanoids were elevated in the CPB group versus the Comparison, including AA (p = 0.008), 15-HETE (p=0.020), 12-HETE (p=0.007), 11-HETE (p=0.009), 8-HETE (p=0.002), 5-HETE (p=0.016), 15-Keto-PGE2 (p=0.008), PGD1 (p=0.006), PGE1 (p=0.030), and 10-HDHA (p=0.023), while DHA remained lower (p<0.001). LTB4 also remained lower in the CPB group post-operatively (p = 0.042). Within the CPB group, 10 plasma eicosanoids increased from pre- to post-surgery. No significant changes in any plasma eicosanoid were observed in the Comparison group across time points.

### Epithelial Barrier Dysfunction and Cytokines

#### Gut EBD

Claudin-2 (p=0.01) and claudin-3 (p=0.006) were elevated in the CPB group compared with the Comparison group at the pre-operative time point, indicating baseline intestinal barrier compromise in CHD patients before any surgical intervention (**Figure 3C**). FABP2 did not differ between groups pre-operatively (p= 0.613). Post-operatively, all three EBD markers were elevated in the CPB group: FABP2 (p<0.0001), claudin-2 (p=0.001), and claudin-3 (p<0.001). Within the CPB group, FABP2 (p<0.0001), claudin-2 (p=0.02), and claudin-3 (p=0.018) were elevated post-surgery compared to pre-surgery. No changes in any EBD marker were observed in the Comparison group between timepoints. The post-operative rise in FABP2, a specific marker of enterocyte injury, is particularly notable, as it directly indicates intestinal epithelial damage attributable to CPB.

#### Cytokines

In the cytokine analysis, Interleukin (IL)-1β (p= 0.036) and Interferon-γ (p= 0.004) were elevated pre-operatively (**Figure 3D**). Post-operatively, multiple cytokines were elevated in the CPB group — IL-1β (p<0.001), Tumor necrosis factor (TNF)-α (p < 0.001), IL-6 (p< 0.0001), IFN-γ (pg/mL, p=0.0002), and IL-8 (p<0.0001) and IL-10 (p<0.0001). The remaining cytokines did not differ from POD #1 through POD #3. Within the CPB group, IL-10, IFN-γ, TNF-α, IL-6, and IL-8 were all increased post-surgery compared to pre-surgery (all p<0.0001), while IL-1β trended toward an increase (p=0.057). Interestingly, IL-10 was decreased on POD #1 (p=0.0002), and IL-6 remained elevated (p=0.0001). In the Comparison group, IL-10 (p<0.001) and IL-6 (p=0.003) were increased post-op.

#### Canonical Correlation Analysis and Mediation Analysis

##### Canonical Correlation Analysis

Regularized canonical correlation analysis (rCCA) was performed to assess multivariate associations between the gut microbiome and host metabolite profiles. Clustered image maps (CIM) revealed organized co-variation blocks between groups of taxa and metabolites (**Figure 4**). At the post-operative timepoint, SCFA-producing species — including *Blautia wexlerae, Blautia obeum*, *Faecalibacterium prausnitzii*, and *Ruminococcus torques* — co-varied with stool butyric acid and propionic acid in the rCCA relevance networks, while Pseudomonadota-associated species clustered with pro-inflammatory eicosanoid metabolites. Plasma cytokine panels showed covariation among IL-6, IL-8, and TNF-α with taxa from the Enterobacteriaceae. Plasma EBD markers (FABP2, claudin-2, claudin-3) showed multivariate associations with microbiome composition, supporting a link between microbial dysbiosis and intestinal EBD.

**Figure 4.**
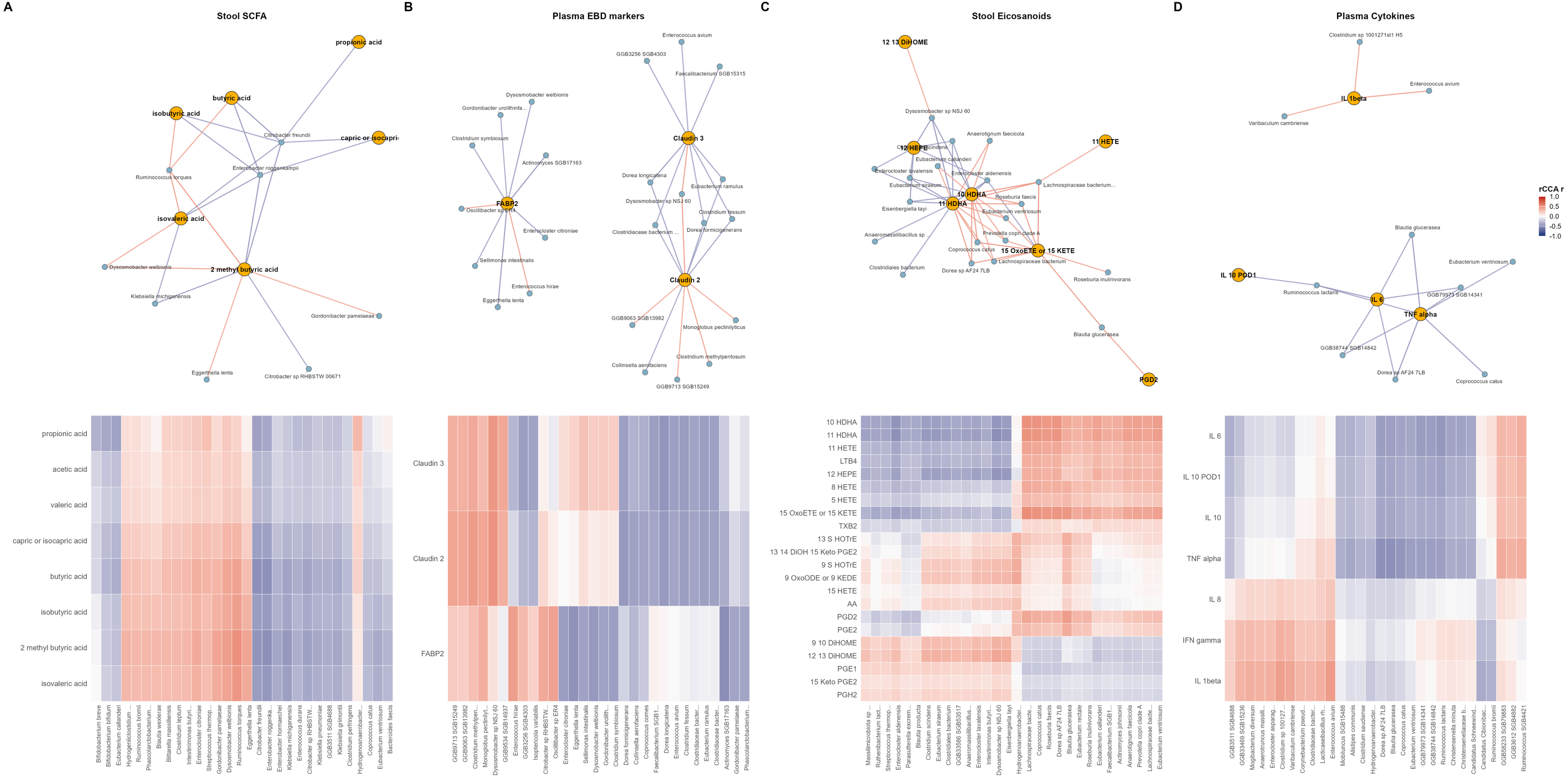
Microbiome-metabolite multivariate associations in CPB patients. Regularized canonical correlation analysis (rCCA) was performed to relate species-level gut microbiome composition to four host metabolite/biomarker panels. **(A)** Stool short-chain fatty acids (SCFA); **(B)** plasma epithelial-barrier-dysfunction (EBD) markers (FABP2, claudin-2, claudin-3); **(C)** stool eicosanoids; **(D)** plasma cytokines. *Top row:* relevance networks showing metabolite-taxon pairs with rCCA correlation magnitude ≥ 0.40 (summed across the first two canonical components). Orange nodes are metabolites/biomarkers; blue nodes are species. Edge color encodes the sign and magnitude of the rCCA correlation (red = positive, blue = negative; scale at right). *Bottom row:* clustered image maps (CIM) of the corresponding rCCA correlation matrices, with row (metabolite) and column (species). Each heatmap shows the 30 species with the largest maximum absolute correlation to any metabolite in that panel. All panels are restricted to CPB patients at the post-operative time point. Analyses of plasma SCFA and plasma eicosanoids are shown in Figure S1.

##### Mediation Analysis

Microbiome mediation analysis using MODIMA was performed to formally test whether the gut microbiome mediates the effect of surgical group (CPB versus Comparison) on downstream metabolomic and inflammatory outcomes at the post-operative timepoint (**Figure 5**). Of the 77 outcomes tested across six panels, four reached nominal significance (all p = 0.045): stool butyric acid, stool isobutyric acid, IL-10, and IL-10 on POD#1 — all within the SCFA and cytokine panels — although none survived BH correction (adjusted p = 0.246). In addition, all three EBD markers trended toward microbiome-mediated effects (FABP2: p = 0.055; claudin-3: p = 0.060; claudin-2: p = 0.074), as did 15 of 22 stool eicosanoids tested (all p = 0.088), stool isovaleric acid (p = 0.069), and plasma isovaleric acid (p = 0.058). The broad, consistent trending signal across the stool eicosanoid panel — in which the exposure-to-microbiome path is shared and only the microbiome-to-response association varies — suggests a common microbially mediated mechanism linking CPB-associated dysbiosis to gut eicosanoid production.

**Figure 5.**
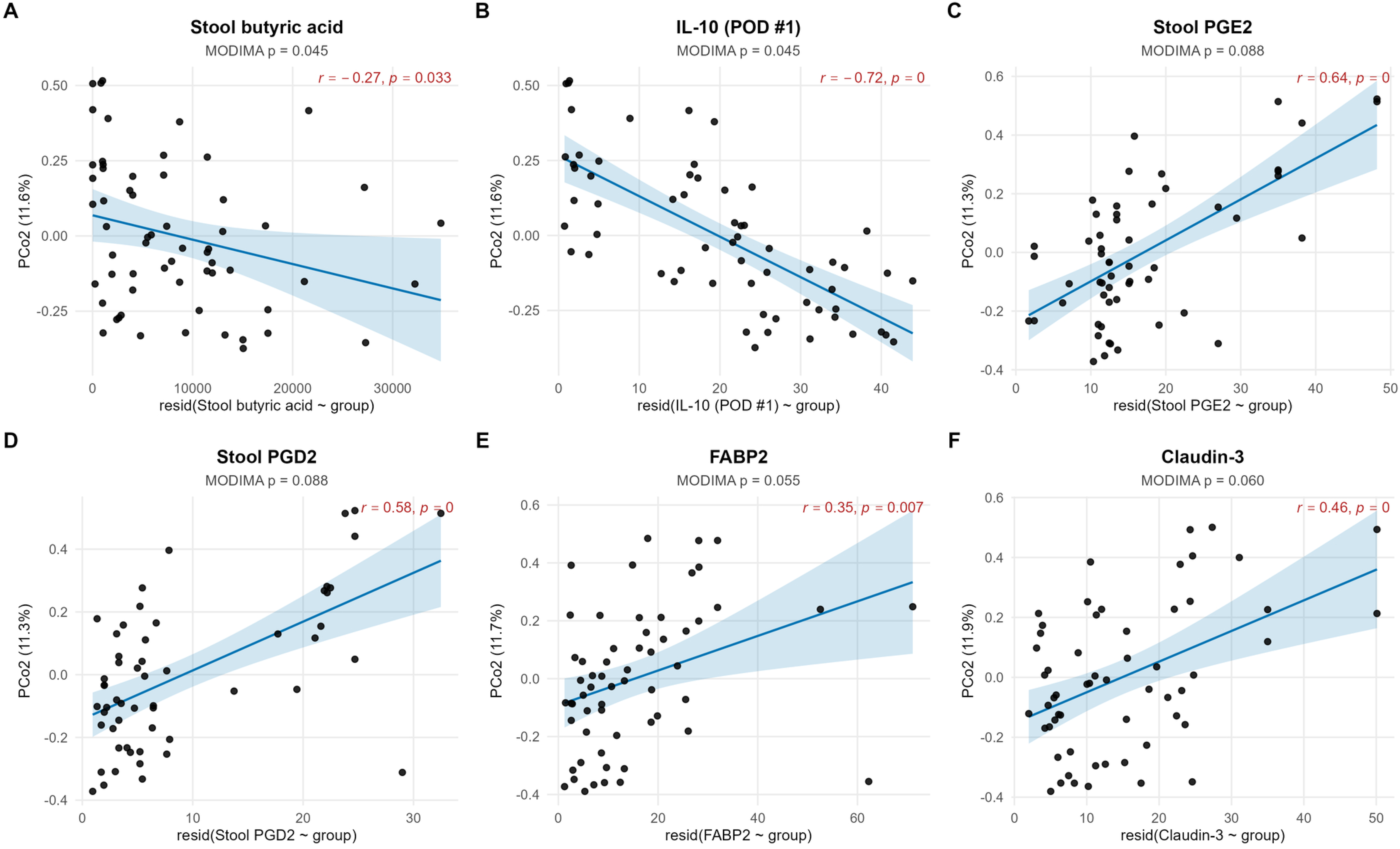
Microbiome-mediated effects of CPB on post-operative metabolites and inflammatory markers. Mediation analysis was performed with the surgical group (Control vs CPB) as the exposure (X), gut microbiome composition as the mediator (M), and each of six post-operative outcomes as the response (Y). Six outcomes are shown for which the omnibus MODIMA test reached p < 0.1: (A) stool butyric acid; (B) plasma IL-10 on post-operative day 1; (C) stool PGE2; (D) stool PGD2; (E) plasma FABP2; (F) plasma claudin-3. Each panel shows the conditional association (Mediator ↔ Response | Exposure): the principal coordinate (PCo) of the microbiome distance matrix after projecting out the exposure effect is plotted on the y-axis against the residual outcome on the x-axis. For each outcome, both PCo1 and PCo2 were tested via Pearson correlation; the PCo2 axis with p < 0.05 is shown. The fitted regression line is shown in blue with a 95% confidence band.

## Discussion

This study defines the gut-heart axis in pediatric congenital heart disease through simultaneous, multi-domain characterization of the gut microbiome, metabolome, epithelial barrier integrity, and systemic cytokine response across the surgical window. The central finding is a two-hit model: children with CHD enter surgery with pre-existing gut dysbiosis, intestinal barrier compromise, and systemic inflammatory priming; cardiopulmonary bypass then delivers a surgery-specific second insult that amplifies each of these abnormalities in a manner not observed after non-cardiac surgery of comparable duration and perioperative management. Together, these findings position the gut as a tractable biological driver of CPB-related morbidity and identify the perioperative microbiome as a candidate therapeutic target in pediatric cardiac surgery.

### Pre-existing Gut Dysbiosis and CPB as a Gut Disruptor

A central and clinically important observation is that gut dysbiosis in CHD precedes surgery. Prior work from our group and others has documented postoperative dysbiosis and barrier dysfunction following pediatric CPB^6,7,18^. Our data demonstrates that *Blautia* depletion and reduced plasma SCFA levels are detectable before the surgical insult, and that claudin-2 and claudin-3 are already elevated pre-operatively with further post-op increases relative to a non-cardiac surgical comparison group, despite similar antibiotic regimens, NPO times, and anesthetic protocols. The elevation of cytokines pre-operatively indicates that this gut-barrier compromise is accompanied by systemic inflammatory priming, establishing heightened immune reactivity prior to surgery. In addition, the CPB pre-operative microbiome exhibited a distinct signature, with enrichment of butanediol biosynthesis and fermentation pathways, associated with a shift towards increased pro-inflammatory

### Enterobacter and Klebsiella

The comparison group showed only two altered microbial metabolic pathways between timepoints, while the CPB group exhibited 30, with 27 enriched post-operatively. Post-operatively, 111 pathways were differentially abundant between groups, 106 enriched in CPB patients. This functional remodeling was not attributed to perioperative exposures. These findings support the conclusion that the gut injury effect of CPB and its associated inflammatory insult is exacerbated by the pro-inflammatory gut milieu. Our data suggest that the microbiome presents an opportunity for intervention prior to surgery. The functional pathway data point to a post-operative shift toward a pathobiont-dominated community. The broad enrichment of biosynthetic pathways in CPB patients — spanning amino acid biosynthesis, heme biosynthesis, assimilatory sulfate reduction, fatty acid β-oxidation, and enterobacterial common antigen biosynthesis — represents a coordinated functional signature of an expanding Enterobacteriaceae population rather than a commensal community.

Consistent with gut oxidative injury, cobalamin biosynthesis was effectively abolished after CPB, paralleling the selective post-operative loss of Methanobrevibacter and other Euryarchaeota, organisms whose extreme oxygen sensitivity makes them markers of a disrupted anaerobic niche^32^. Most importantly, lipid IVA biosynthesis — a committed step in the synthesis of the lipopolysaccharide endotoxin — was enriched in post-operative CPB patients. This raises the possibility that the expanded pathobiont community has increased capacity for endotoxin production; direct measurement of luminal and circulating endotoxin in future studies will be needed to determine whether this pathway enrichment translates to increased endotoxin translocation across the compromised barrier^33^.

### Convergent Metabolomic Disruption: SCFA Depletion and Eicosanoid Excess

The metabolomic data reveal two parallel, directionally opposing changes that together characterize the post-CPB gut-systemic state: a collapse in protective short-chain fatty acid availability and a surge in pro-inflammatory eicosanoid production. SCFAs — particularly butyrate and propionate — are the primary energy substrates for colonocytes^34^, suppress NF-κB-driven inflammatory gene expression through histone deacetylase inhibition, strengthen tight junction protein assembly, and promote peripheral regulatory T-cell differentiation^35-39^. Their depletion removes protective signaling from the intestinal epithelium during acute surgical stress. Notably, the SCFA deficit was not mirrored by a loss of encoded pathway potential; HUMAnN-derived biosynthetic pathways were instead enriched post-operatively, reflecting the gene-dense facultative anaerobes that expand after CPB. This indicates that the SCFA deficit reflects the taxonomic loss of specific obligate anaerobe producers (Blautia, Roseburia, Faecalibacterium, Lachnospiraceae) and their fermentative flux, rather than a global loss of encoded pathway capacity, although we cannot exclude that altered substrate availability or post-operative changes in luminal conditions also contribute to the gap between encoded pathway capacity and measured SCFA output.

The concurrent elevation of gut and plasma eicosanoids represents the equal and opposite metabolomic pressure. Eicosanoids are potent lipid mediators of inflammatory signaling: prostanoids activate immune cell recruitment and vascular permeability, HETEs amplify neutrophil activation and leukotriene biosynthesis, and oxidized linoleic acid metabolites such as 9(S)-HOTrE and 9-OxoODE serve as bioactive ligands for PPAR-γ and other immune transcription factors^40^. The elevated stool eicosanoid levels before surgery indicate that the gut inflammatory tone is already chronically elevated in CHD, consistent with the SCFA deficit removing the anti-inflammatory constraint on gut immune activation. The post-operative surge represents a robust amplification of this pre-existing state by the CPB insult, consistent with enhanced arachidonic acid release from epithelial and immune cell membranes under oxidative and ischemic stress^41^.

### Barrier Compromise and Microbial Mechanistic Mediators

The dissociation between claudin and FABP2 behavior across the two time points encodes a mechanistically informative two-stage model of gut barrier failure that distinguishes chronic structural vulnerability from acute CPB-specific enterocyte injury. Pre-operative claudin-2 and claudin-3 elevation in the absence of FABP2 elevation reflects paracellular permeability through disrupted tight junction architecture — a marker of chronic barrier remodeling^38,42^ consistent with the SCFA deficit and gut eicosanoid excess already present before surgery. SCFA depletion directly weakens tight junctions by reducing butyrate-driven AMPK activation and claudin protein expression. The post-operative rise in FABP2 indicates acute cellular disruption in epithelial cells themselves^6,43^. Prior studies of gut injury after adult CPB demonstrated similar postoperative FABP2 elevation, linking it to the ischemic phase of bypass and subsequent endotoxemi^33^. Our data extend this to the pediatric CHD population and demonstrate its absence in the comparison group — confirming specificity to cardiac surgery and CPB.

The canonical correlation and mediation analyses provide two complementary layers of statistical evidence linking the gut microbiome to the metabolomic and inflammatory changes observed in this study. These analyses address a key inferential gap that exists when microbiome, metabolomic, and inflammatory data are examined in isolation: they establish whether the co-occurring abnormalities are structured (CCA) and whether the microbiome occupies a causal position in the exposure-to-outcome pathway (MODIMA), rather than being a correlated parallel process. The convergence of CCA co-variation structure with MODIMA mediation signals — both centering on the SCFA–commensal and eicosanoid–pathobiont axes — provides a coherent inferential framework that positions the gut microbiome as a mechanistic intermediary, rather than a passive bystander, in the inflammatory cascade triggered by CPB. However, we note that although the convergent CCA and MODIMA signals are consistent with a microbiome-mediated causal model, the observational design cannot rule out the possibility that CHD-related hemodynamic or metabolic alterations drive dysbiosis and barrier compromise in parallel rather than in series; interventional studies that manipulate the pre-operative microbiome will be required to establish directionality.

The IL-10 trajectory merits particular attention within this framework. IL-10 was among the outcomes nominally mediated through the microbiome and demonstrated a surge-and-collapse pattern. The IL-10 rises sharply post-operatively alongside pro-inflammatory cytokines, consistent with an initial compensatory anti-inflammatory response, but then falls on POD#1 even as IL-6 remains elevated. This exhaustion of the regulatory cytokine response — occurring while the pro-inflammatory axis persists — may represent a critical inflection point at which the balance tips toward sustained systemic inflammation^11^, and its association with microbiome composition in the MODIMA analysis suggests that the post-operative microbial community may influence the durability of this anti-inflammatory counter-regulation^44^.

### Clinical Implications and Study Limitations

The findings of this study have direct implications for the clinical management of children with CHD undergoing cardiac surgery. The demonstration that gut dysbiosis and barrier compromise precede surgery by a measurable and characterizable degree identifies the pre-operative period as a biologically rational window for intervention that has not previously been defined in pediatric CHD. Therapeutic strategies that could plausibly improve the pre-operative gut ecosystem include prebiotic supplementation to promote SCFA-producing commensals and probiotic administration with organisms such as *Lactobacillus* and *Bifidobacterium* species, which are known to support barrier integrity^45^. This has previously been shown in animal models of cardiopulmonary bypass and in adult cardiac surgery^18,46^ and has been explored as a therapeutic in adult cardiac patients^47^, but has not been evaluated in the pediatric CHD population.

There are several limitations of this study. The sample size is modest, and the power for some analyses is constrained. This study is observational, and interventional studies are required to establish true causality. Additionally, this study does not incorporate a direct control group, but rather a comparison group to account for perioperative variables. Identifying a group of children with CHD undergoing non-cardiac surgery that is also non-GI surgery is challenging, given the low frequency of these surgeries. This limits the direct effects of the inflammatory gut milieu, thereby exacerbating CPB-induced gut injury. Future studies that include multiple sites to develop a true control group of non-cardiac surgery CHD patients are warranted. The microbiome is not static and undergoes distinct changes during the first few years of life, when the majority of cardiac surgeries occur; this is a confounding variable that this study was not powered to account for. Larger studies with subgroup analyses in larger populations are necessary to understand differences and implications across age groups. Additionally, the cyanotic status of the patients also presents challenges with alterations in gut perfusion and degrees of hypoxemia, which can alter the microbial composition and baseline gut inflammation pre-operatively.

### Conclusions

This study demonstrates that children with congenital heart disease undergoing cardiac surgery with cardiopulmonary bypass enter the operating room with a pre-existing dysbiotic gut ecosystem characterized by depleted microbial diversity, impaired SCFA-producing capacity, elevated gut eicosanoid production, intestinal tight junction disruption, and systemic cytokine activation. Cardiopulmonary bypass delivers a surgery-specific second insult that amplifies each of these abnormalities, depleting SCFA-producing commensals and reorganizing microbial functional potential toward a pathobiont-associated, endotoxin-generating profile, broadening eicosanoid excess into the systemic circulation, inducing direct enterocyte injury, and overwhelming the anti-inflammatory regulatory cytokine response. These findings provide a scientific foundation for prospective trials of perioperative microbiome-directed interventions in this high-risk population and set the stage for a companion analysis examining subgroup-specific biology and preoperative predictive signatures in this patient population.

## Data Availability

The datasets generated and/or analyzed during the current study are available from the corresponding author on reasonable request.

## Abbreviations

CHD: Congenital Heart Disease
CPB: Cardiopulmonary Bypass
EBD: Epithelial Barrier Dysfunction
SCFA: Short-Chain Fatty Acid
FABP2: Fatty Acid Binding Protein
PG: Prostaglandin
IL: Interleukin

## Acknowledgements

Funding for this study was supported by American Heart Association – 23CDA1045271, the National Institute for General Medical Sciences - P20GM103427-19, and the Child Health Research Institute. We also would like to acknowledge the Clinical Pharmacology Laboratory for the metabolomic quantification and the Bioinformatics Team at the University of Nebraska-Lincoln. We would like to thank the CHRI for assistance with study coordination and grant and manuscript writing review. Special thanks to Sarah O’Neill for her tireless dedication to this study.

---

**Figure S1. Microbiome-metabolite multivariate associations in CPB patients.** Regularized canonical correlation analysis (rCCA) was performed to relate genus-level gut microbiome composition to two host metabolite/biomarker panels. **(A)** Plasma SCFA; **(B)** Plasma eicosanoids. *Top row:* relevance networks showing metabolite-taxon pairs with rCCA correlation magnitude ≥ 0.40 (summed across the first two canonical components). Orange nodes are metabolites/biomarkers; blue nodes are the genus. Edge color encodes the sign and magnitude of the rCCA correlation (red = positive, blue = negative; scale at right). *Bottom row:* clustered image maps (CIM) of the corresponding rCCA correlation matrices, with row (metabolite) and column (genus). Each heatmap shows the 30 genus with the largest maximum absolute correlation to any metabolite in that panel. All panels are restricted to CPB patients at the post-operative time point.

